# Effect of Vitamin D_3_ supplementation vs. dietary-hygienic measures on SARS-COV-2 infection rates in hospital workers with 25-hydroxyvitamin D3 [25(OH)D3] levels ≥20 ng/mL

**DOI:** 10.1101/2022.07.12.22277450

**Authors:** Maria Elena Romero-Ibarguengoitia, Dalia Gutiérrez-González, Carlos Cantú-López, Miguel Angel Sanz-Sánchez, Arnulfo González-Cantú

**Affiliations:** Department of Research, Hospital Clínica Nova; San Nicolás de los Garza, N.L., Mexico; Universidad de Monterrey, Vicerrectoría de Ciencias de la Salud, Departamento de Ciencias Clínicas, San Pedro Garza García, N.L. Mexico; General Management, Hospital Clínica Nova; San Nicolás de los Garza, N.L., Mexico; Department of Endocrinology, Hospital Clínica Nova; San Nicolás de los Garza, N.L., Mexico

**Author notes:** Corresponding Author: Arnulfo González-Cantú, *Hospital Clínica Nova*, Av. del Bosque 139, Cuauhtémoc, 66450 San Nicolás de los Garza, N.L., México.

**Keywords:** SARS-COV-2, vitamin D_3_, supplementation

## Abstract

**Background:** There is scant information on the effect of supplementation with vitamin D_3_ in SARS-COV-2 infection cases when patient 25-hydroxyvitamin D_3_ [25(OH)D3] levels are between 20-100ng/mL. Our aim was to evaluate the effect of supplementation with vitamin D_3_ vs. dietary-hygienic measures on the SARS-COV-2 infection rate in participants with serum 25(OH)D_3_ levels ≥20ng/mL.

**Methods:** We invited hospital workers with 25(OH)D_3_ levels between 20-100 ng/mL and no previous SARS-COV-2 infection; they were randomized as follows: treatment options were a) vitamin D_3_ supplementation (52,000 IU monthly, G1) or b) dietary-hygienic measures (G2). We conducted a 3- to 6-month follow-up of SARS-COV-2 infections. Participants with 25(OH)D_3_ levels <20 ng/mL were also analyzed. We divided these latter participants depending on whether they were supplemented (G3) or not (G4).

**Results:** We analyzed 198 participants, with an average age of 44.4 (SD 9) years, and 130 (65.7%) were women. G1 had less cases of SARS-COV-2 infection than G2 after a follow-up of 3- to 6-months (p<0.05). There were no differences between G3 and G4 at the 3- and 6-month follow-up cutoff points (p>0.05). Using mixed effect Cox regression analysis in 164 participants that completed six months of follow-up, vitamin D_3_ supplementation appeared to act as a protective factor against SARS-COV-2 infection (HR 0.21, p=0.008) in G1 and G2. None of the participants treated with the supplementation doses had serum 25(OH)D_3_ levels > 100ng/mL.

**Conclusion:** Vitamin D_3_ supplementation in participants with 25(OH)D_3_ levels between 20-100 ng/mL have a lower rate of SARS-COV-2 infection in comparison with the use of dietary-hygienic measures at six months follow-up.

## Introduction

COVID-19 is an infectious disease caused by the newly discovered coronavirus SARS-CoV-2; its clinical spectrum ranges from asymptomatic infection to critical and fatal illness^1,2^ [1,2]. The first case was reported in Wuhan, China, in December 2019, while the first case in Latin America was detected in Brazil in February 2020 ^3,4^ [3,4]. As of March 13, 2022, approximately 458,479,635 cases were confirmed, and 3 million deaths were reported worldwide. Furthermore,10,712,423,741 vaccine doses have been applied^5^ [5].

To date, prevention remains the cornerstone of management to decrease infection rates. In late 2020, the authorization of SARS-CoV-2 emergency vaccines led to partial pandemic control. However, further studies will be key to obtain clear evidence on its treatment and prevention^6^ [6].

There is controversy on the use of vitamin D_3_ supplementation in the prevention of SARS-CoV-2^7^ [7]. A meta-analysis concluded that a low serum 25-hydroxyvitamin D_3_ [25(OH)D_3_] level was significantly associated with a higher risk of SARS-CoV-2 infection ^8^ [8]. However, there is limited data on the link between SARS-COV-2 infection and vitamin D_3_ supplementation in individuals with normal 25(OH)D_3_ levels.

The rationale for this study was to establish a relationship between vitamin D_3_ supplementation and the incidence of SARS-CoV-2 infection in a prospective study. The target population included health workers at high risk of SARS-COV-2 infection and vitamin D serum values ≥ 20 ng/mL.

We expected a positive effect of vitamin D supplementation on the immune system (innate and adaptive immunity) ^9,10^ [9,10], and that the group who underwent supplementation would develop fewer SARS-COV-2 cases.

The study aimed to evaluate the effect of vitamin D_3_ supplementation vs. dietary-hygienic measures on the development of SARS-COV-2 infection in participants with serum 25(OH)D_3_ ≥20 ng/mL (primary outcome). Secondarily, we compared a group of hospital workers with serum 25(OH)D_3_ < 20 ng/mL that could not or had not been supplemented with vitamin D_3_.

We hypothesized that low serum vitamin D3 is associated with an increased risk of SARS-COV-2 infection. We believed that we would find a difference in the development of SARS-COV-2 in participants with serum 25(OH)D_3_ ≥20 ng/mL who received vitamin D_3_ supplementation vs. dietary-hygienic measures ^11^[11].

## Methods and Materials

The study was a prospective, quasi-experimental study that followed the CONSORT guidelines^25^ [21]. It included health workers from a hospital in Northern Mexico, the *Hospital Clínica Nova* (HCN), at a northern latitude of 25°45’ and western latitude of 100°17‵. This hospital was converted to a COVID-19 Hospital in March 2020. Participant recruitment began in May-August 2020, when the initial baseline serum vitamin D level was obtained. Participant follow-up continued from August 2020 through January 2021.

The study was conducted per The Code of Ethics of the World Medical Association (Declaration of Helsinki) for experiments in humans. It was also approved by the local IRB of the *Universidad de Monterrey* (Ref. 30062020-a-CN-CI) and registered with the clinical trial number NCT04810949 (first released 23/03/2021). Due to the study’s nature, an informed consent form was signed by each participant, and two witnesses. The author, AGC, enrolled the participants and assigned them to each intervention.

The inclusion criteria were age between 18 and 65 years old, both genders, absence of infection by SARS-COV-2, influenza H1N1, influenza A or B at the time of serum vitamin D_3_ determination, absence of infection at any site (bacteria or fungi), and participants had to be hospital workers at the HCN. In addition, participants were excluded if their serum 25(OH)D_3_ was >100 ng/mL, if they had previously received supplements containing vitamin D_3_, and if they were pregnant.

Individuals were invited to participate in the study after signing the consent form. They were directed to the laboratory to provide serum samples to determine their levels of 25(OH)D_3_. We used the Elecsys total vitamin D test with COBAS 6000 (e601) equipment (Wiesbaden Germany). The CV was 4.1%, the analytic specificity for 25(OH)D_3_ was 100%, and the analytic sensibility was established at 4.01ng/mL. Calibration of the equipment was performed by the laboratory staff every time there was a change in the lot, and the calibration curve factor was 1 (goal 0.8- 1.2).

Based on the results, we classified participants into four groups. The first and second groups had 25(OH)D_3_ >20 ng/mL and they were randomized in a 1:1 ratio; the participants had two treatment options: the first, was supplementation with vitamin D_3_ 52,000 IU in a single dose, monthly, for three months (13 tablets of 4,000 UI, G1). The total dose is based on the Endocrine Society’s Guidelines^26,27^ [26,27]; however, we decided to administer it every month instead of a daily, since some of this frequency’s benefits have been discussed elsewere^28^[25]. The second option was treatment based on dietary-hygienic measures (G2), such as sun exposure for at least 10 minutes per day between 10:00-18:00 hrs., and foods that were rich in vitamin D_3_ and D_2_ (fish, meat, eggs, milk, mushrooms, and, almonds)^29,30^[29,30].

The third and fourth groups had 25(OH)D_3_ levels ≤20 ng/mL. These groups were not part of the randomization process, since we considered that all subjects with vitamin D_3_ <20ng/mL should receive vitamin D_3_ supplementation^31^ [31], and they were referred to their personal primary care physician (different primary care physician) for treatment and follow-up (G3). Nevertheless, a certain number of participants decided, of their own volition, not to receive vitamin D_3_ supplementation (G4), but since they were health workers and had a medical record from the same hospital where the study was conducted, we could still follow them over time. Our secondary aim was to also determine the incidence of SARS-COV-2 infection in this group. We, therefore, had 4 groups for comparisons.

Variables obtained from the medical record were age, gender, occupation, diabetes mellitus, hypertension, allergies, asthma, smoking history, previous hospitalizations, and BMI. The participants were monitored monthly during follow-up, every month by telephone, and every 3 months in a face-to-face interview. We inquired on COVID-19 symptoms (myalgias, hyposmia, cough, malaise and fatigue, nasal congestion, fever, diarrhea, thoracic pain, shaking chills, nausea, and vomiting), and whether they had been diagnosed with SARS-COV-2 infection by serologic or swab tests (PCR). Also, every month patients were asked if they had been consuming food with vitamin D_3_ or D_2_ using a 24h food recall questionary. Serum 25(OH)D_3_ was measured at baseline and after three months of follow-up.

The relative risks that could be present in the different groups were increased serum vitamin D levels (> 100ng /ml) in G1 and decreased vitamin D serum levels < 20ng/mL in G2. The risks in G3 and G4 could not be controlled by the research group.

The relative risks that could be present in the different groups were an increase in vitamin D serum levels > 100ng /ml in G1, and decreased vitamin D serum levels < 20ng/mL in G2. The risk in G3 and G4 could not be controlled by the research group.

## Statistical analysis

Two researchers reviewed the quality control of the database and anonymized it. Normality assumption was evaluated with the Shapiro Wilk test and frequency histograms. Descriptive statistics such as the mean, the standard deviation for quantitative variables, frequencies, and percentages for categorical variables, were computed. Chi-square tests and ANOVA were used to compare the categorical and quantitative data between groups. Kaplan-Meier curves and the Log-rank test were used to evaluate the difference in SARS-COV-2 infection between groups. We conducted a mixed effect Cox regression analysis in the groups with 25(OH)D_3_ ≥20 ng/mL and <20 ng/mL, in which the dependent variable was SARS-COV-2 infection at six months. The covariates computed in the model were vitamin D_3_ supplementation, age, and gender. The seasonal variation was a random effect. Missing data were handled by complete case analysis. For the simple randomization, we used random number generation with a binomial distribution, and a probability of 50%. The author, AGC, generated the randomization sequence.

The statistical programs used were SPSS version 25 (IBM, USA) and R software version 4.0.3 (R Core Team, Vienna Austria). The analysis was two-tailed. A p-value < 0.05 was considered statistically significant. The sample size included all hospital workers that agreed to provide serum samples for 25(OH)D_3_ testing.

## Results

Initially, 205 hospital workers were considered for the study; five had to be excluded because they had previously contracted SARS-COV-2, before the study started. Additionally, two individuals in the G1 and G2 withdrew their consent to participate.

In the end, 198 participants were analyzed. These participants were distributed in four groups based on their 25(OH)D_3_ levels and the type of treatment administered. Group 1 (G1) included participants with 25(OH)D_3_ level ≥20ng/mL plus supplementation (52,000IU/month) (n=43). Group 2 (G2) consisted of participants with 25(OH)D_3_ level ≥20ng/mL managed with dietary-hygienic measures (n=42). Group 3 (G3) had participants with serum 25(OH)D_3_ <20ng/mL plus supplementation (90,000IU/month, average dose provided by their treating physician) (n=28). Finally, group 4 (G4) included participants with 25(OH)D_3_ <20ng/mL, without supplementation (n=85).

The mean (SD) age in the four groups was 44.4 (9.1) years (p>0.05), with no difference between groups, and 130 were female (65.7%). The three most frequent professions in all the groups were: physician, nurses, and administrative workers, and no difference was established among G1-G4 (p>0.05). There were also no differences in terms of smoking history, Body Mass Index (BMI), allergies, Type 2 diabetes, hypertension, and asthma (p>0.05). Demographic data and medical history of the groups are described in Table 1.

**Table 1.** Demographics and Medical History

| Groups n=198 | G1 | G2 | p-value | G3 | G4 |  |
| --- | --- | --- | --- | --- | --- | --- |
|  | n= 43 | n= 42 |  | n= 28 | n= 85 | p-value* |
| Female | 26 (60) | 19 (45) | >0.05 | 20 (71) | 65 (76) | >0.05 |
|  | Profession |  |  |  |  |  |
| Physician (%) | 15 (35) | 16 (38) | >0.05 | 7(25) | 23(27) | >0.05 |
| Nurse (%) | 19(44) | 15(36) |  | 13(46) | 39(46) |  |
| Dentist (%) | 1(2) | 2(5) |  | 1(4) | 7(8) |  |
| Maintenance (%) | 2(5) | 3(7) |  | 0 | 1(1) |  |
| Administrative (%) | 5(12) | 1(2) |  | 4(14) | 9(11) |  |
| Nutritionist (%) | 0 | 3(7) |  | 2(7) | 2(2) |  |
| Other (%) | 1(2) | 2(5) |  | 1(4) | 4(5) |  |
|  | Medical History |  |  |  |  |  |
| Tobacco(%) | 1(2) | 4(10) | >0.05 | 2(7) | 3(4) | >0.05 |
| BMI (SD) | 26.4(5.1) | 26.3(3.5) | >0.05 | 27.8(2.1) | 28.2(4.5) | >0.05 |
| Type 2 Diabetes (%) | 5(11) | 1(2) | >0.05 | 2(7) | 6(7) | >0.05 |
| Allergies (%) | 7(16) | 7(17) | >0.05 | 7(25) | 20(24) | >0.05 |
| Hypertension (%) | 6(14) | 3(7.5) | >0.05 | 5(18) | 11(13) | >0.05 |
| Asthma (%) | 2(5) | 2(5) | >0.05 | 0 | 0 |  |

\*Chi-square test was performed for between-group comparisons. A p-value $\leq 0.05$ was considered statistically significant. Parentheses represent frequency or standard deviation. Abbreviation: SD: standard deviation; BMI: body mass index; G1: 25(OH)D3 $\geq 20$ ng/mL with
supplementation; G2: 25(OH)D3 ≥ 20 ng/mL without supplementation; G3: 25(OH)D3 <20 ng/mL with supplementation; G4 25(OH)D3 <20 ng/mL without supplementation.

The mean (SD) serum baseline 25(OH)D_3_ levels reported per group were: 27.1 (6.7) ng/mL in G1, 26.6 (5.5) ng/mL in G2, 15.4 (3.4) ng/mL in G3, and 14.9 (3.1) ng/mL in G4, p<0.001. Serum levels of 25(OH)D_3_ were again measured after three months of follow-up. The values were as follows: G1 33.8 (7.1) ng/mL, G2 22.4 (6.9) ng/mL, G3 38.2 (8.5) ng/mL and G4 22.1 (5.8) ng/mL, p<0.001.

At the 3-month cutoff, 51 (25.8%) of the 198 workers had developed SARS-COV-2 infection (naïve variant). The distribution of cases by groups was: G2 14 (33%) and G1 3 (7%), p= 0.002; G4 27 (32%) and G3 7 (25%), p>0.05. When comparing between the four groups there was a statistical difference, p=0.017.

One hundred eighty-seven (187) individuals were followed for four months; of these, 56 (29.4%) developed SARS-COV-2 infection (naïve variant). The proportion of SARS-COV-2 infection cases by groups was: G2 14 (34%) and G1 6 (14%), p= 0.002; G4 29 (38%), and G3 7 (26%), p = 0.04. When comparing between the four groups there was a statistical difference, p=0.041 Additionally, we studied 167 workers that completed a 6-month follow-up. Not all participants completed this 6-month follow-up because they were recruited at a later date, the SARS-COV-2 vaccination campaign had begun, and we considered that vaccination could affect our results; we therefore completed our study follow-up, and they were not included in this timeline analysis. However, during that period, 56 (33.5%) developed Covid-19 (naïve variant). The number of cases by group was as follows: G2 13 (39%) and G1 5 (13%), p<0.001; G4 29 (40%) and G3 9 (37%), p >0.05. When comparing between the four groups there was statistical difference, p=0.031. Figure 1 shows comparisons between G1 vs. G2 and G3 vs G4. The Nursing Department had the highest proportion of SARS-COV-2 infections, although no statistical difference between groups was detected (p> 0.05). The most frequently reported symptoms were myalgias and fatigue. A total of 8 (14%) participants were hospitalized during the study, with no statistical difference between groups (p>0.05). Only one worker in the G4 group required intubation. There were no deaths in the study. Data on the SARS-COV-2 infections during the 6-month follow-up is presented in Table 2.

**Table 2.** SARS-COV-2 infection at 6-month follow-up

| n=167 | G1<br>(n=38) | G2<br>(n=33) | p-value | G3<br>(n=24) | G4<br>(n=72) | p-value |
| --- | --- | --- | --- | --- | --- | --- |
| SARS-COV-2 | 5 ( 13) | 13 (39) | 0.01 | 9 (37) | 29 (40) | >0.05 |
| Physician (%) | 2(15) | 3(23) | >0.05 | 2(22) | 5(17) | >0.05 |
| Nurse(%) | 2(11) | 8(61) | 0.003 | 6(67) | 17(59) | >0.05 |
| Dentist (%) | 0 | 0 |  | 0 | 3(10) |  |
| Administrative (%) | 1(20) | 1(100) | >0.05 | 0 | 3(10) |  |
| Nutritionist (%) | 0 | 1(33.) |  | 1(11) | 1(3) | >0.05 |
| SARS-COV-2 associated symptoms |  |  |  |  |  |  |
| Dry cough (%) | 0(0) | 0(0) |  | 2(22) | 5(17) | >0.05 |
| Fatigue (%) | 2(40%) | 1(7.7%) | >0.05 | 4(44) | 4(14) | 0.049 |
| Sore throat (%) | 1(20%) | 0 |  | 1(11) | 5(17) | >0.05 |
| Nasal congestion (%) | 1(20%) | 1(7.7%) | >0.05 | 0 | 2(7) |  |
| Fever (%) | 1(20%) | 2(15.4%) | >0.05 | 1(11) | 2(7) | >0.05 |
| Myalgias (%) | 4(80) | 2(8) | 0.009 | 4(44) | 4(13.8%) | 0.049 |
| Diarrhea (%) | 1(20) | 1(8) | >0.05 | 1(11) | 0 |  |
| Chest pain (%) | 1(20) | 0 |  | 0 | 2(7) |  |
| Shaking chills (%) | 1(20) | 0 |  | 1(11) | 0 |  |
| Nausea (%) | 1(20) | 0 |  | 0 | 0 |  |
| Hyposmia (%) | 0 | 1(8) |  |  | 0 |  |
| Hospitalizations (%) | 2 (40) | 1 (8) | >0.05 | 3 (33) | 2 (7) | 0.04 |
| Intubations (%) | 0 (0) | 0 (0) |  | 0 | 1 (3) |  |
| Reservoir mask (%) | 1(20) | 0(0) |  | 0 | 0 |  |
| Oxygen via nasal prongs (%) | 0 (0) | 0 (0) |  | 0 | 1 (3) |  |
| Pneumonia (%) | 3(60) | 1(7.7) | 0.017 | 1(11) | 2(7) | >0.05 |

A p-value ≤ 0.05 was considered statistically significant. Abbreviation: SD: standard deviation; G1: 25(OH)D3 ≥ 20 ng/mL with supplementation; G2: 25(OH)D3 ≥ 20 ng/mL without supplementation; G3: 25(OH)D3 <20 ng/mL with supplementation; G4: 25(OH)D3 <20 ng/mL without supplementation. Parentheses represent frequency or standard deviation

**Figure legends.**
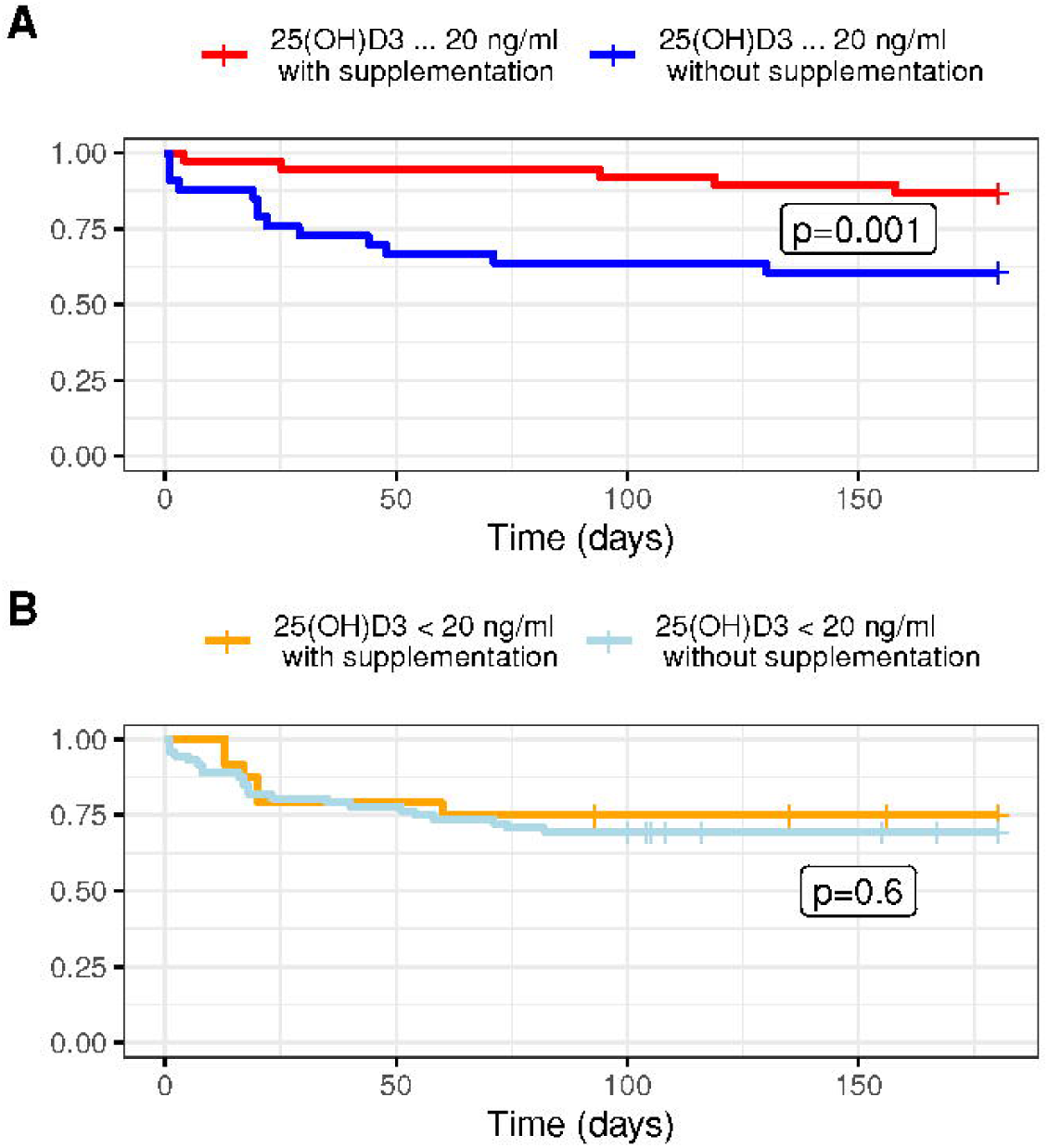
SARS-COV-2-free survival. Kaplan Meier curves of the SARS-CoV-2 infection rate during the six-month follow-up. The images compare the 4 groups according to their baseline vitamin status and vitamin D3. Figure A shows the group with serum 25(OH)D3 ≥20 ng/mL. Figure B shows 25(OH)D3 ≤20 ng/mL The lowest rate of SARS-COV-2 infection occurred in the group with vitamin D > 20ng/mL plus supplementation for three months, 52000IU per month.

We conducted a mixed effect Cox regression analysis with the participants who were followed for six months (n=164). The final covariates computed in the model were vitamin D_3_ supplementation, age, and gender. The seasonal variation was a random effect. In the Group ≥ 20ng/mL 25(OH)D_3_, the resulting HR was 0.21 (SE 0.58, p= 0.008) for vitamin D_3_ supplementation; age, 0.97 (SE 0.26, p=0.25); and gender, 0.85 (SE 0.49, p=0.76). In the Group < 20ng/mL 25(OH)D_3_, the HR was 1.15 (SE 00.39, p= 0.72) for vitamin D_3_ supplementation; age, 1.001 (SE 0.02, p=0.94); and gender, 0.52 (SE 0.48, p=0.18). Other covariates that were explored were BMI, Type 2 diabetes, and hypertension, but they were not statistically significant, so they were eliminated from the final models.

In the G1 and G3 groups that were supplemented with 52,000 IU/month and 90,000IU for three months, respectively, none of the participants had 25(OH)D_3_ levels >100 ng/mL and no adverse events were reported.

## Discussion

This study demonstrated that vitamin D_3_ supplementation for three months led to a decrease in the rate of SARS-COV-2 infection in the group of participants with 25(OH)D_3_ levels ≥20 ng/mL throughout the 3–6-month follow-up when compared with dietary-hygienic measures. In this study, participants with 25(OH)D_3_ levels < 20 ng/mL had a higher rate of SARS-COV-2 infection; however, upon comparison of groups (G3 and G4), the supplemented group had less frequent SARS-COV-2 infections at four months of follow-up but not at six months.

Previous studies, including a metanalysis, revealed an association of low serum 25(OH)D_3_ levels with SARS-COV-2 infection^8,12^[8,12]. Another study reported a correlation of this deficiency with an aging population, aside from associated comorbidities such as diabetes mellitus, hypertension, or obesity^12,13^ [12,13]. This suggests that there is an inverse relation between SARS-COV-2 infection and 25(OH)D_3_ levels. As mentioned previously, we did find an association between low levels of 25(OH)D_3_ and SARS-COV-2 infection; however, when we evaluated the comorbidities, we did not find a difference in participants with comorbidities. Thus, the most important predictor of infection in the regression models was the supplementation with vitamin D_3_ and low 25(OH)D_3_ values.

A study conducted in Barcelona demonstrated that participants that had previously received vitamin D for four months were at a decreased risk of acquiring SARS-COV-2 infection (HR = 0.95, CI 0.91–0.98), but this was not applicable in individuals on calcifediol^14^ [14]. A previous clinical trial conducted in Mexico City included health workers who were randomized to receive either 4000 IU of vitamin D_3_ for 30 days or placebo and were followed for 45 days. The results showed that independently of the baseline 25(OH)D_3_ (that in the study was mostly deficient), the supplemented group had a lower incidence of SARS-COV-2 infection^15^ [15].

A study conducted in Veteran patients in the US Defense Department health system, showed an inverse dose-response relationship between continuously increasing 25(OH)D concentrations (from 15 to 60 ng/mL with supplementation), and a parallel decreased probability of requiring COVID-19-related hospitalization^16^ [16].

As previously stated, our study supports the importance of supplementation. Nevertheless, there is still a lack of information to be obtained in clinical trials on SARS-COV-2 management, and on the potential benefit of supplementation with vitamin D_3_ in participants with 25(OH)D3 ≥20 ng/mL to decrease SARS-COV-2 infection risk; therein lies the value of our study in which we also evaluated the effect in this group of participants in a prospectively manner.

We observed that in our group of participants who followed dietary-hygienic measures, their 25(OH)D_3_ levels had decreased when measured a second time. This may result from the difficulty among the participants to closely adhere to the recommendations provided by the physician, such as sun exposure, work hours, and diet. Further, it is important to note that the second measurement of 25(OH)D_3_ was obtained in the winter season. These factors could explain the increased number of cases of SARS-COV-2 infection and underscore the usefulness of supplementation in the population during this time of the year. Our regression model that adjusted the seasonal variation, reinforce the importance of supplementation.

Since there is evidence of a greater risk of acquiring a SARS-COV-2 infection and an increase in its severity among participants with 25(OH)D_3_ levels <20 ng/mL ^**8**,**17–19**^[8,15,17], we considered it would not be ethical to randomize treatment in these groups of participants. They were referred to the primary care physician to initiate supplementation. Unfortunately, despite the treatment options and recommendations, some participants decided not to follow them. However, the supplemented group had a lower incidence of SARS-COV-2 at four months of follow-up but not at six months. We believe that there was an important effect of 3-month supplementation but that was not sustained for six months, so future studies must be conducted were a sustained supplementation is evaluated.

From a mechanistic point of view, vitamin D status could also be an index of nitric oxide concentrations induced by solar UVA rays and these may act in concert to potentially prevent the COVID-19-dependent cytokine storm and induced inflammation^20,21^ [20,21]. Also, vitamin D could protect against viral infection through the innate immune response. The induction of cathelicidin and defensins can block viral entry to the cell and suppress viral replication. Another mechanism is by promoting autophagy of the virus, expressing autophagy marker LC3, downregulating the mTOR pathway, promoting Beclin 1and PI3KC3, and stimulating the formation of autophagosomes to indirectly facilitate viral clearance. Therefore, vitamin D could have an important role in maintaining the balance between autophagy and apoptosis, and thus maximize the antiviral response to infection^10,22^ [10,18].

Vitamin D is also a regulator of the adaptive immune response by inducing regulatory T cells that are critical to the induction of immune tolerance, and play a role in preventing the cytokine storm associated with severe respiratory disease caused by viral infections. Vitamin D, via its active metabolite 1,25(OH)_2_D, limits the maturation of dendritic cells, and hence, their ability to present antigen to T cells, and change the T cell profile from the proinflammatory Th1 and Th17 to Th2 and T regulatory subsets, that inhibit the proinflammatory processes^10^ [10].

Activation of the vitamin D receptor could play a modulatory role to the host responses in the acute respiratory distress syndrome by decreasing cytokines, producing a shift toward amplified adaptive Th2 immune responses, regulating the renin-angiotensin-bradykinin system, modulating neutrophil activity, and maintaining the integrity of the pulmonary epithelial barrier, thus promoting epithelial repair, and decreasing the coagulability and prothrombotic tendency associated with SARS-COV-2 infection ^22–24^ [18-20].

One of our study’s limitations is the sample size in each group; however, using a formula for the proportion difference between independent groups of 26%, we achieved a power of 86%. Also, some participants did not complete their follow-up at 4- and 6-months because of SARS-COV-2 vaccination. We decided to discontinue the study once patients were vaccinated since it could affect our primary outcome. Finally, the study was conducted in only one center and mostly in the winter, so a multicenter study conducted during all seasons is required in the future, and with a greater sample size to confirm the risks and benefits of supplementation in participants with 25(OH)D_3_ levels <20 ng/mL, in terms of the acquisition of SARS-COV-2 infections.

The risks of the protocol were minimal since the supplementation of vitamin D_3_ at doses of 52,000 units per month is not related to any adverse event. Vitamin D hypervitaminosis occurs when serum levels are > 100ng / ml. The administered supplementation dose had a very low risk of causing hypervitaminosis D. Nevertheless, participants’ serum levels were monitored at three months as a security measure. Another risk of the study was the possibility of an allergic reaction to any component of the vitamin D_3_ presentation formula, but none of the participants developed this condition.

## Data Availability

All data produced in the present study are available upon reasonable request to the authors

## Acknowledgments

We are grateful to Rosalinda González Facio, Laura Patricia Montelongo, and Gerardo Del Río Parra for their assistance in participant follow-up during the trial, and their review of medical records. Likewise, we appreciate the Nursing Department’s coordination of appointments.

## Author Contribution

Conceptualization: MERI, AGC, DGG, and MASS. Formal analysis: MERI, AGC, DGG, and CCG. Investigation: MERI, AGC, DGG, MASS, and CCG. Resources: MASS, Writing – original draft: MERI, AGC, DGG, MASS, and CCG. Writing – review and editing: MERI, AGC. Project administration: MERI, DGG, and MASS. Supervision: MERI, AGC, and MASS. Funding acquisition: MASS. All authors contributed to this article and approved the submitted version.

## Data Availability

Data are available upon reasonable request to the corresponding author

## Ethics Statement

This research project was approved by the local IRB of the *Universidad de Monterrey* (Ref. 30062020-a-CN-CI). An informed consent form was obtained from each participant.

## Author Approval

all authors read an approved the final version of the manuscript.

## Protocol access

This study protocol can be reviewed in Clinicaltrials.gov.

